# SARS-CoV-2 seropositivity after infection and antibody response to mRNA-based vaccination

**DOI:** 10.1101/2021.02.09.21251319

**Authors:** Emily J. Ciccone, Deanna R. Zhu, Rawan Ajeen, Evans K. Lodge, Bonnie E. Shook-Sa, Ross M. Boyce, Allison E. Aiello, COVID HCP Study Team

**Author notes:** **Corresponding Author Information:** Gillings School of Global Public Health, CB #7435, Chapel Hill, NC 27599, USA.

## Abstract

The effect of SARS-CoV-2 infection on response to mRNA-based SARS-CoV-2 vaccines is not well-described. We assessed longitudinal SARS-CoV-2-specific antibody responses pre- and post-vaccination among individuals with and without prior infection. The antibody response to the first vaccine dose was almost two-fold higher in individuals who were seropositive before vaccination compared to those who were seronegative, suggesting that prior infection primes the immune response to the first dose of mRNA-based vaccine.

---

The impact of prior SARS-CoV-2 infection on antibody response to mRNA-based vaccines has not yet been well-characterized. The Centers for Disease Control and Prevention (CDC) recently suggested that individuals with documented immunity might defer vaccination for 90 days to expedite access for those without immunity, but data supporting these guidelines are limited.^1^ Specifically, there are few studies describing the response to vaccination among previously infected individuals. Early reports suggest that prior infection may prime the immune response to the first vaccine dose, but these reports did not assess longitudinal changes in antibody levels before vaccination.^2–4^ This information is important as the timing of natural infection and fluctuations in immunity over time may impact the vaccine response.^5,6^ Here, we examined longitudinal SARS-CoV-2 antibody responses among healthcare personnel (HCP) with and without evidence of prior infection to examine the effect of natural infection on vaccination response.

We conducted a longitudinal observational cohort study of HCP at a large academic medical center, which began enrolling in July 2020. Its purpose was to examine risk factors for infection and identify changes in antibodies to SARS-CoV-2 over time.^7^ The study was approved by the University of North Carolina Institutional Review Board (20-0942), and informed consent was obtained from all participants. Enrolled individuals provided blood samples every two weeks for the first 12 weeks and monthly thereafter. mRNA-based SARS-CoV-2 vaccinations (Pfizer-BioNTech [BNT162b2] and Moderna [mRNA-1273]) were available to HCP from the insitution starting in mid-December 2020 or earlier through vaccine trials. For all pre- and post-vaccination samples, we measured total immunoglobulin antibodies specific to the receptor binding domain of the SARS-CoV-2 spike protein by ELISA as previously described.^8^ We used a positive to negative ratio (P/N) cut-off of 2.57 to ensure 99.5% specificity per CDC recommendations for SARS-CoV-2 serology assays.^9^ Exact Wilcoxon rank sum tests with corresponding 95% confidence intervals (CI)^10^ were used to compare median P/N levels between seropositive and seronegative participants and change in P/N for those with paired pre- and post-vaccination measurements.

As of February 17, 2021, the study has enrolled 213 HCP with 194 (91%) providing at least one blood sample. One individual was excluded from this analysis due to co-enrollment in a SARS-CoV-2 treatment trial. Table 1 presents the cohort demographics. Five participants (3%) had antibodies to SARS-CoV-2 at their baseline study visit, herein “prevalent seropositive” (Figure 1a). Notably, one demonstrated persistent seropositivity for over six months, while another remained seropositive for more than three months before reverting to seronegative one month prior to vaccination. Five participants (3%) became seropositive during the study, herein “incident seropositive” (Figure 1a); four of those five individuals (80%) reported COVID-19 symptoms prior to seroconversion.

**Table 1.** Demographics, serostatus, and vaccinations for the total study population and the sub-sample of participants with pre- and post-vaccination antibody measurements.

| **Characteristics** | **Total study population (n=193)** | **Participants with pre- and post-vaccine antibody results (n=93)<sup>a</sup>** |
| --- | --- | --- |
| **Age, median (IQR)** | 38 (31-44) | 39 (31-46) |
| **Female sex, n (%)** | 132 (68) | 63 (68) |
| **Race/Ethnicity, n (%)** |  |  |
| *Black/Non-Hispanic* | 8 (4) | 2 (2) |
| *Asian/Non-Hispanic* | 11 (6) | 5 (5) |
| *White/Non-Hispanic* | 138 (72) | 71 (76) |
| *Other or 2 or more races/Non-Hispanic* | 10 (5) | 6 (6) |
| *Unidentified/Non-Hispanic, Unknown* | 16 (8) | 7 (8) |
| *Hispanic Ethnicity (of any race)* | 10 (5) | 2 (2) |
| **Serostatus prior to vaccination, n (%)** |  |  |
| *Seronegative* | 183 (96) | 87 (94) |
| *Seropositive at study enrollment* | 5 (3) | 4 (4) |
| *Incident seropositive* | 5 (3) | 2 (2) |
| **Vaccinations received to date, n (%)** |  |  |
| *None* | 6 (3) | N/A |
| *1 dose* | 6 (3) | 2 (2) |
| *2 doses* | 181 (94) | 91 (98) |

<sup>a</sup>Post-vaccine refers to after at least one dose

**Figure 1.**
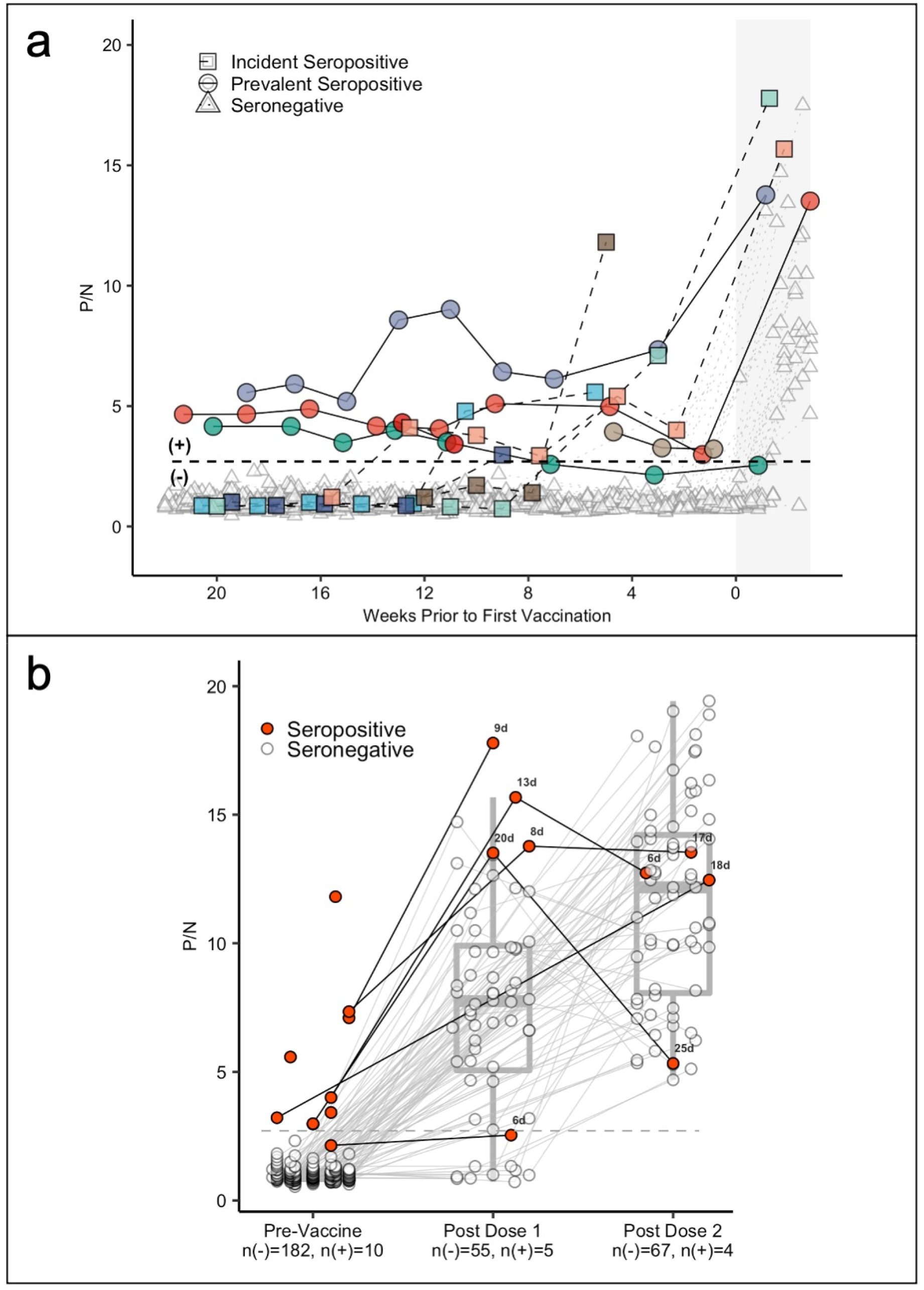
Antibody levels of healthcare personnel over time (a) and in response to vaccination (b). Panel (a) demonstrates antibody levels over time in SARS-CoV-2 seronegative (triangles) and seropositive individuals, including those who were seropositive at enrollment (prevalent seropositive, circles) and those who became seropositive during follow-up (incident seropositive, squares). The dotted line is a P/N ratio of 2.57, the cut-off associated with 99.5% specificity (SARS-CoV-2 Ig-positive above the line, Ig-negative below). The x-axis represents weeks to first vaccine dose; values post-vaccine 1 are shaded. Panel (b) shows box plots with the medians and interquartile ranges of P/N ratios pre-vaccination, post-vaccine 1, and post-vaccine 2 by serostatus (seronegative: n(-), seropositive: n(+)).The upper and lower whiskers extend to the largest and smallest values no further than 1.5 times IQR from the hinge, respectively. Data beyond the end of the whiskers are outliers. For the pre-vaccine time point, the most recent antibody level prior to vaccination (for those who were vaccinated) or most recent antibody level overall (for those who were not vaccinated) is shown. For the post-vaccine time points, the first measurement after 5 days post-vaccination is included. The black numbers next to the circles indicate the number of days between vaccination and sample collection for seropositive individuals (V_1_: median 9, range 6-20; V_2_: median 17.5, range 6-25; Supplemental Figure 1). For seronegative participants, the median time between the first vaccine dose and sample collection was 16 days (range 6 to 34 days); between the second vaccine dose and sample collection it was 17 days (range 6 to 33; Supplemental Figure 1).

Most participants (97%, 187/193) received at least one dose of a SARS-CoV-2 mRNA-based vaccine as part of the institution’s vaccination program or through a vaccine trial. Antibody measurements after at least one vaccine dose were available for 60% (6/10) of seropositive and 48% (87/183) of seronegative participants. One individual tested positive for SARS-CoV-2 by PCR shortly after the second vaccine dose (V_2_); post-V_2_ results were excluded for this participant. We compared antibody responses to the first vaccine dose between participants who were seropositive (at any point) and those who were seronegative prior to vaccination (Figure 1b). Figure 1b shows that seropositive individuals developed an almost two-fold higher antibody response to their first vaccine dose (median P/N=13.8) compared to those who were seronegative (median P/N= 7.6; difference estimate 6.53 (95% CI: 1.6-10.4), Wilcoxon test statistic (W) = 227, p=0.014). Additionally, the post-V_1_ antibody response in seropositive participants was of similar magnitude to the post-V_2_ response among seronegative participants (post-V_1_ seropositive median P/N 13.8 vs. post-V_2_ seronegative 12.2; difference estimate 1.68 (95% CI: −3.3-6.0), W=202, p=0.462). The absolute magnitude of the increase in antibody levels from pre-vaccination to post-V_1_ was greater for seropositive than seronegative participants (median change in P/N from pre to post-V_1_ seropositive 10.5 vs. seronegative 6.4; difference estimate 2.66 (95% CI: −1.6-5.8), W=181, p=0.22).

Our findings provide preliminary evidence that prior SARS-CoV-2 infection may impact the immune response to the first dose of mRNA-based SARS-CoV-2 vaccine. Specifically, we observed that the response to one vaccine dose among seropositive individuals was almost two-fold higher than that of seronegative individuals and of similar magnitude to the response to two vaccine doses among seronegative individuals.

Our study is limited by a relatively smaller sample size of seropositive compared to seronegative participants and should therefore be replicated by future work. However, we noted a significant difference in response to the first vaccination despite the small sample size, and our results corroborate recent non-peer-reviewed findings.^2–4^ We also extend previous work by demonstrating that the observed immune respone to vaccination among seropositive individuals is robust across varying patterns of antibody responses over time and regardless of timing of past infection, which ranged from 1 to 6 months prior to vaccination.

In conclusion, prior SARS-CoV-2 infection appears to prime the immune response to the first dose of mRNA-based vaccine. Further research on the durability of the response to the first dose of SARS-CoV-2 vaccine among previously infected individuals could help guide the allocation of the limited supply of mRNA-based vaccines. As 27 million Americans have been infected with SARS-CoV-2 to date,^11^ the potential for some individuals to forgo a second vaccination may have substantial impact on vaccine distribution strategies.

## Data Availability

The study described in this manuscript is ongoing, and therefore, the data are not yet available to others. Once study follow-up is completed, the data will be made available by request.

## Acknowledgements

We thank the participants for their willingness to contribute to advancing our understanding of the SARS-CoV-2 epidemic and its impact on healthcare personnel, especially during the early and uncertain months of the pandemic. We also appreciate the hard work of the graduate student assistants from the University of North Carolina Gillings School of Global Public Health (Elle Law, BS, Elyse Miller, BA, and Paul Zivich, MPH) and the phlebomists who contributed to the sample collection, protocols, and day-to-day implementation of the study (these individuals received compensation for their work on the study). We acknowledge Premkumar Lakshmanane, PhD who spearheaded development of the ELISA assay and produced and supplied the necessary antigens for the study through funding from the National Cancer Institute [1U54CA260543]. Finally, we thank Subhashini Sellers, MD, MSc and Meghan Rebuli, PhD (University of North Carolina School of Medicine) and Bailey Fosdick, PhD (Colorado State University Department of Statistics) for their support of this study and its analysis. These contributors did not receive any contribution for their role in the study.

## Author Contribution Statement

EJC designed and supervised the study, composed and revised the paper, and helped to secure study funding.

DRZ contributed to study design, led the data analysis, and assisted with generation of the figures.

RA assisted with data collection and analysis and revised the paper.

EKL contributed to study design, and assisted with generation of the figures.\

BES assisted with data analysis and revision of the paper.

RMB conceptualized, designed, and supervised the study, revised the paper, and helped to secure study funding.

AEA conceptualized, designed, and supervised the study, composed and revised the paper, and helped to secure study funding.

All authors, including those in the COVID HCP Study Team, have participated in preparing the manuscript and have reviewed and approved its final, submitted version.

