## Supplemental Figure 1 for "SARS-CoV-2 seropositivity after infection and antibody response to mRNA-based vaccination"

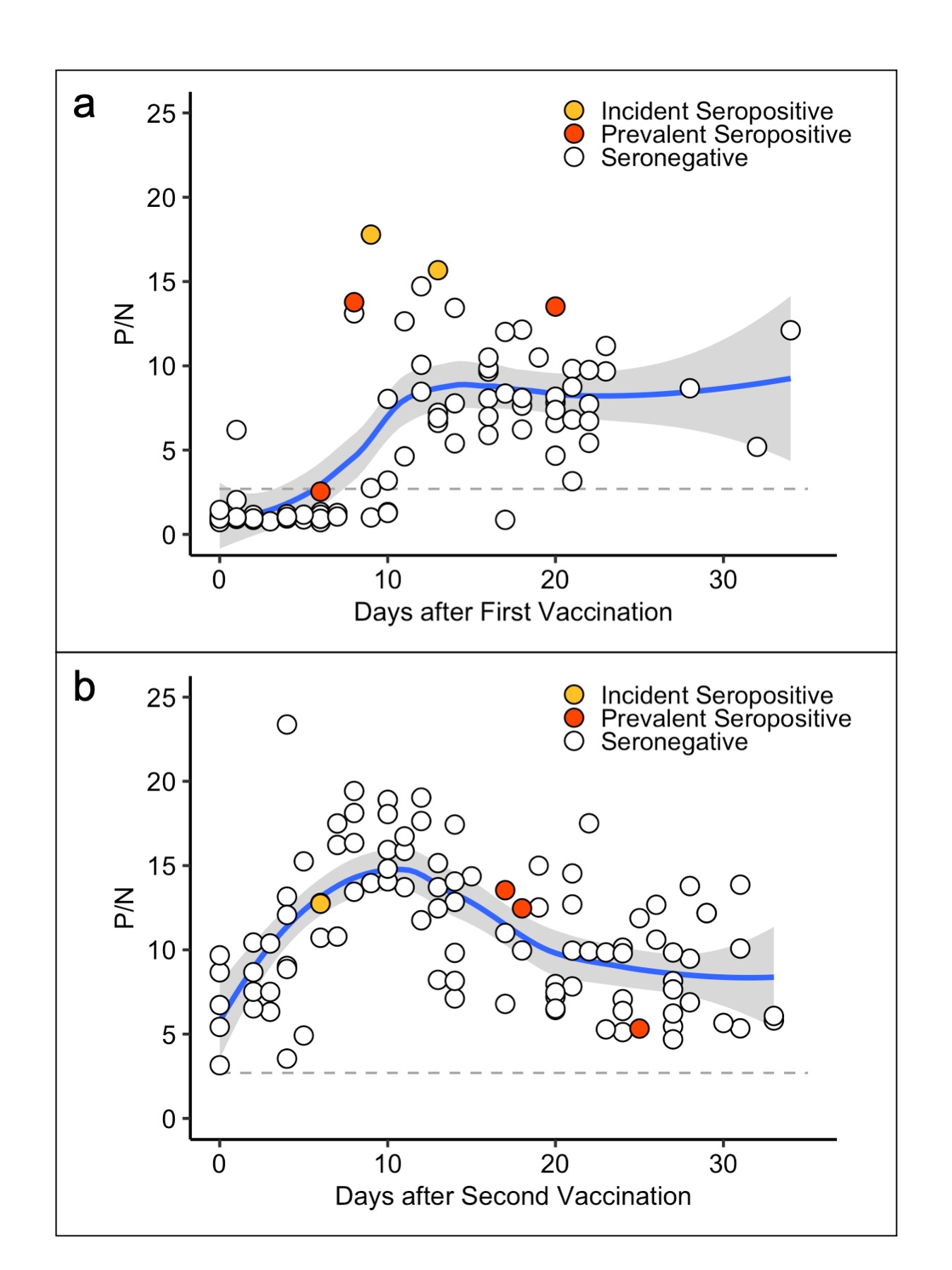
**Supplementary Figure 1. Antibody responses to SARS-CoV-2 after first (a) and second (b) vaccination**. Panel (a) shows SARS-CoV-2 Ig P/N values after the first dose of SARS-CoV-2 mRNA-based vaccine. All samples collected between the date of first vaccination up to date of second vaccination were included. Panel b shows antibody levels after the second dose of vaccine. All study visits occurring within 40 days of second vaccination were included.

Dotted lines represent a P/N ratio of 2.57, the cut-off associated with 99.5% specificity (SARS-CoV-2 Ig-positive above the line, Ig-negative below). Blue lines represent Loess regression with associated shaded 95% confidence intervals in gray.
